# Added diagnostic accuracy after [18F]flutemetamol PET scanning in young patients with dementia in an academic memory clinic setting

**DOI:** 10.1101/2021.11.30.21266815

**Authors:** Ron L.H. Handels, Marissa D. Zwan, Wiesje Pelkmans, Geert Jan Biessels, Peter van Domburg, Erik Hoff, Gerwin Roks, Niki Schoonenboom, Niek Verweij, Bart N.M. van Berckel, Wiesje M. van der Flier, Frans Verhey, Philip Scheltens, Femke H. Bouwman

**Author notes:** Below we provide a draft description of results of one of the research questions as set in the Flutemetamol study (trial registry link https://www.clinicaltrialsregister.eu/ctr-search/trial/2012-002303-18/NL). During the project the validity of the method was discussed, in light of the swiftly developing state of the art in the field. After consultation of an external expert* the decision was made not to finalize the manuscript and refrain from submitting the work to a journal. Prof. Dr. Frisoni from Memory Clinic, Geneva University and University Hospitals, Geneva, Switzerland.

## Abstract

**Introduction:** A timely diagnosis of Alzheimer’s Disease (AD) in an early stage of dementia is important to support timely access to treatment, advice, and care. The aim of this study was to estimate the diagnostic accuracy of [18F]flutemetamol PET in addition to the usual diagnostic workup for the diagnosis of AD in a memory clinic population with young onset dementia by means of a panel reference-based etiology diagnosis.

**Methods:** in an academic memory clinic early onset dementia cohort (n=211) the nosological diagnosis was set by usual diagnostic workup and after including [18F]flutemetamol amyloid PET in a stepwise approach. To assess the change in proportion correctly diagnosed, the diagnosis with and without [18F]flutemetamol PET was related to a panel-based reference standard, serving as gold standard, consisting of 3 neurologists who relied on available clinical information over 2-year follow-up (n=152; blinded for PET).

**Results:** The panel majority nosology was set as a reference diagnosis in 122 participants, leaving 30 (20%) participants with no majority reached. In 107 (88%) cases post-PET was in line with the reference, and in 103 (84%) the pre-PET diagnosis was in line with the reference. The difference was 3.3% (95% CI -3.5% to 10.1%; p=0.424).

**Discussion:** [18F]flutemetamol PET did not significantly improve the diagnostic accuracy in young patients with dementia in an academic memory clinic setting. The secondary analyses provided several indications for future research in a narrower subsample of persons with (very) high diagnostic uncertainty and to assess patient relevant health outcomes.

## INTRODUCTION

A timely diagnosis of Alzheimer’s Disease (AD) in an early stage of dementia is considered important to support timely access to treatment, advice, and care [ADI report 2011; Dubois, 2016].

In the last decades diagnostic research has focused on the use of amyloid biomarkers in CSF and on PET to identify amyloid pathology in dementia as well as pre-dementia stages of mild or subjective cognitive impairment, which has led to the development of new clinical and research criteria [Jack, 2018; Albert, 2011; McKhann, 2011].

Fluorine-18 labeled amyloid tracers, among which [18F]flutemetamol with a relatively long half-life, enabled a wide spread application of this amyloid biomarker to serve clinics outside its tracer synthesis facility as compared to for example the [11C] Pittsburg Compound-B (PIB) tracer.

Johnson et al. [2016] indicated the clinical use of amyloid PET is appropriate when the possible diagnosis of AD is uncertain after a comprehensive evaluation by a dementia expert and knowledge of the presence or absence of Amyloid pathology is expected to increase diagnostic certainty and alter management. In addition, recommendations to use and reimburse amyloid PET should rely on its clinical utility in terms of empirical evidence on health outcomes and evidence on cost-effectiveness of the test-treat or test-care pathway [Lijmer, 2009a].

In terms of health outcomes, systematic reviews on the effect of early identification of AD hallmarks did not find empirical evidence of health benefits derived from persons with and without early identified AD genetic profile or pathological hallmarks, but nevertheless reported on potential benefits and harms of early AD testing across the clinical stages of pre-clinical, MCI and dementia in the absence of (disease-modifying) treatment [Dubois, 2016; Paulsen, 2013; ADI, 2011].

In terms of diagnostic accuracy, ideally, a gold standard is available to determine the target condition. However, as this is not the case for AD [Scheltens, 2011] a next best alternative is to rely on a clinical reference standard. A variety of reference standards have been used to compare the diagnostic performance of the clinical practice diagnostic workup without and with using the results from PET imaging. Some studies evaluated amyloid PET in terms of progression from MCI to clinically established AD-type dementia after 2 years, using clinical assessment [Ossenkoppele, 2012] or statistical modelling [Vos, 2015]. Some studies used neuropathology at autopsy [Curtis, 2015; Sanchez-Juan, 2014]. Various studies have not used a reference standard and focused on the proportion of change in clinical diagnosis, mean change in diagnostic certainty and/or proportion clinical (treatment) management before and after disclosing the PET result using various [18F] amyloid tracers (florbetapir, flutemetamol, florbetaben) [Ceccaldi, 2018; Leuzy, 2018; Zwan, 2017; Rabinovici, 2019; Trivino, 2019]. Although this evidence is highly valuable and supports diagnostic test development, a reference standard consisting of conversion from MCI to dementia is mainly limited to reflecting the prognostic value of PET. A reference standard consisting of pathology is limited by disregarding the clinical expression of the underlying cause. A focus on diagnostic change rather than comparing to a reference standard is limited by relying on the assumption that any change is a correct change.

In absence of a gold standard, the use of a panel has been recommended in order to combine multiple test results and construct a reference standard outcome [Rutjes, 2007].

To the best of our knowledge only Hellwig et al. [2018] assessed Amyloid PET in terms of its relation to a clinically relevant reference standard. They used [11C]PIB amyloid PET in 84 patients with a major neurocognitive disorder of uncertain etiology in an academic memory clinic. An etiology diagnosis was set before and after interpreting PET in addition to relevant clinical information (neuropsychological assessment, MRI/CT, CSF). The reference standard consisted of an interdisciplinary board consensus on etiology relying on all available clinical information on clinical symptoms, neuropsychological testing, additional imaging and CSF (from pre-PET and mean 2.4 year follow-up), as well as baseline [11C]PIB and [18F]FDG PET information.

With no longitudinal evidence available on [18F]flutemetamol PET the aim of this study was to estimate the diagnostic accuracy of [18F]flutemetamol PET in addition to the usual practice diagnostic workup for the clinical diagnosis in a memory clinic population of patients with young onset dementia by means of a panel reference-based etiology diagnosis that uses clinical follow-up information. We hypothesize a larger proportion of correctly diagnosed participants after PET compared to the diagnosis before PET, according to the reference standard.

## METHODS

### PARTICIPANTS

In the Dutch Flutemetamol Study, participants received an [18F]flutemetamol PET scan and were followed up for 2 years in a longitudinal prospective cohort design. Participants were recruited between 2012 and 2015 from the VU University Medical Center (n=200) and Maastricht University Medical Centre+ (n=11) if they were suspected of dementia, aged ≤ 70 and mentally competent (MMSE≥18) and diagnostic certainty between 50-90% (see supplemental 1 and Zwan et al. [2017] for details). Written informed consent was obtained from all participants. This study was approved by the medical ethics review committee of the VU University Medical Center (reference number 2012/302).

Participants were omitted from the analysis due to insufficient follow-up information on cognition, behavior and activities of daily living (ADL) (either obtained from test results or from (telephone) interview), or on imaging or biomarker information (see figure 1 for reasons of drop-out).

**Figure 1:**
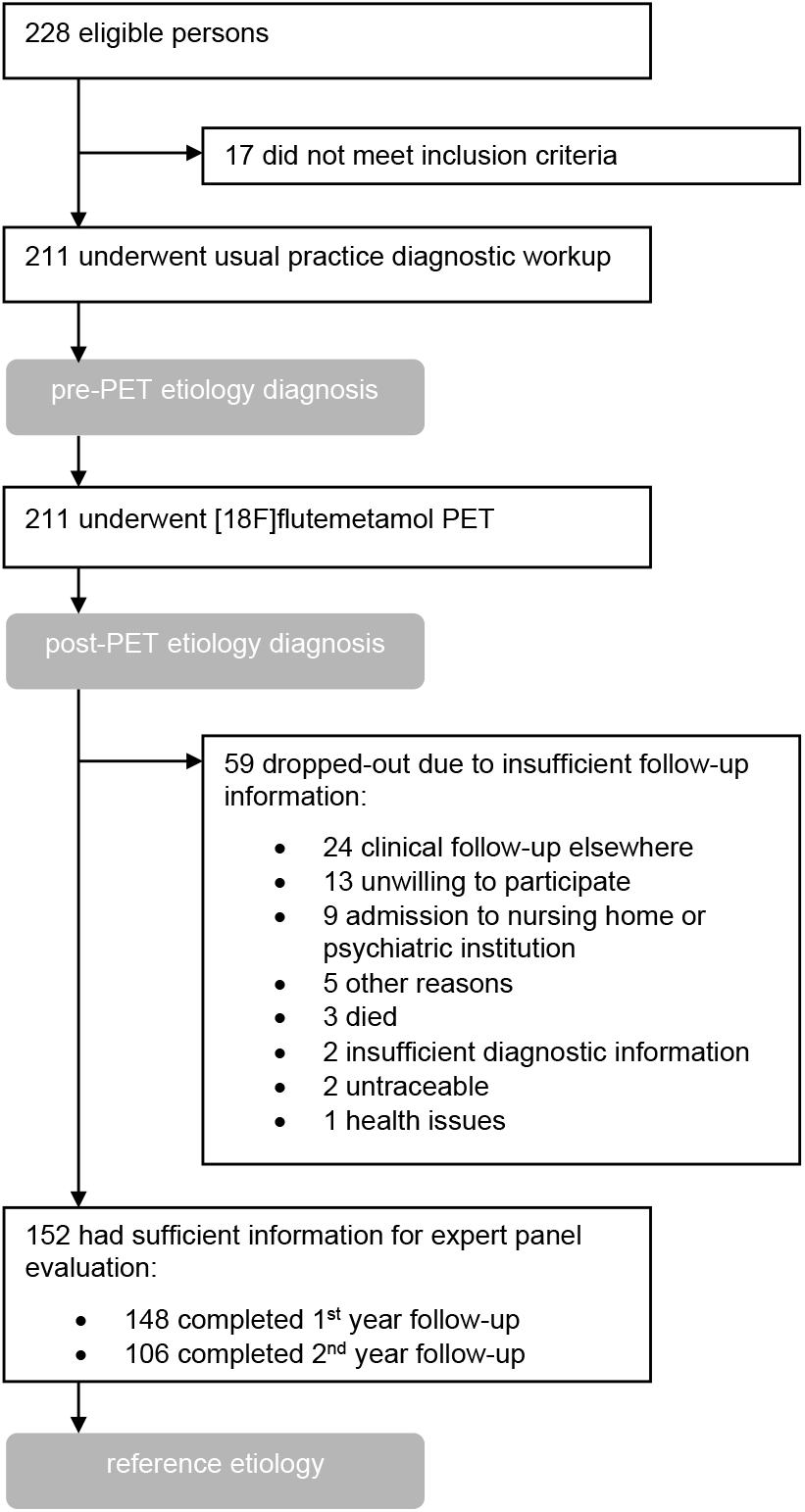
Flowchart of patient inclusion, follow-up and diagnostic accuracy design.

### CLINICAL ASSESSMENTS

At baseline, participants received a standardized clinical dementia evaluation including information on demographics, medical history, informant-based history, physical examination, neurological examination, neuropsychological examinations, screening laboratory tests, brain MRI, and neuropsychological testing [van der Flier, 2014; Aalten, 2014]. In the absence of contraindications, a lumbar puncture was performed and additional investigations considered relevant (e.g. FDG-PET, psychiatric assessment, blood analysis). A standardized clinical follow-up was performed after 1 and 2 years, at which the same information was obtained except from imaging, CSF and blood samples.

If a person was not able to attend the memory clinic for an assessment, a telephone interview was performed to obtain as much information as possible on the presence of dementia-related symptoms and progression.

See supplemental 2 for an example vignette with hypothetical diagnostic information.

### [18F]flutemetamol PET

In both centers, [18F]flutemetamol PET scans were made on a Gemini TF-64 PET/CT scanner (Philips Medical Systems, Best, the Netherlands) [Surti, 2007]. Ninety minutes after a bolus injection of 191 ± 10 MBq [18F]flutemetamol, patients underwent a low-dose CT scan followed by a 20-minute (i.e., 4 frames of 5 minutes) PET scan. Scans were checked for movement and frames were summed to obtain a static (20-minute) image for each patient (except for one patient in whom the last frame was not used due to extensive head movement). Scans were visually assessed and dichotomously rated as either amyloid-positive or amyloid-negative by the local nuclear medicine physician, who completed the training program for visual interpretation of [18F]flutemetamol images. Readers were blinded to clinical information, except for brain MRI.

### PRE- AND POST-PET ETIOLOGY DIAGNOSIS

At baseline, an etiology diagnosis was set in a multidisciplinary meeting according to established criteria [McKhann, 2011; Rascovsky, 2011; Roman, 1993; McKeith, 2005; Boeve, 2003; Litvan, 1996] based on the diagnostic information obtained in usual practice, except for altering the usual practice by blinding them for CSF results (further referred to as pre-PET etiology diagnosis).

After about 4-12 weeks a [18F]flutemetamol PET scan was performed and rated as AD or non-AD by a specialist in nuclear medicine. The treating neurologist combined this information with the pre-PET diagnostic information to set an etiology diagnosis, guided by AD diagnostic research criteria [McKhann, 2011] (further referred to as post-PET etiology diagnosis).

For the purpose of this study, lumbar puncture results were not disclosed before the impact of PET results had been assessed. All etiology diagnoses were rated as AD, vascular dementia (VAD), frontotemporal dementia (FTD), Dementia with Lewy bodies (DLB), other neurodegenerative disease (other NDD) and no neurodegenerative disease (no NDD) as well as their certainty, using standardized forms (see supplemental 3).

### REFERENCE ETIOLOGICAL DIAGNOSIS

A similar method to Handels et al. [2016] was followed and recommendations for designing a panel diagnosis [Bertens, 2013; Handels, 2014] were taken into account as much as possible.

All information described in the paragraph ‘clinical assessments’ was summarized on a digital vignette, which was rated online independently by 3 neurologists (among GB, PD, EH, GR, NS and NV) with over [x] years of clinical experience operating in a local (PD, EH, GR, NS and NV) or university hospital (GB). Ratings were done blinded for [18F]flutemetamol PET ensuring no diagnostic review bias. They individually rated for each etiology (AD, VAD, FTD, DLB, otherNDD, noNDD) if it determined the clinical picture on a 15-point scale (see supplemental 3). Each panelist’s set of ratings was standardized by dividing it by his/her total attributed points to all etiologies. If at least 2 panelists had identified the same etiology or identical combination of etiologies with a standardized rating of ≥0.33 (i.e. one third of the total), this was set as the majority etiology (see supplemental 4 for examples). If not, it was set as inconclusive. Alternatively, the panelists’ ratings of the inconclusive cases were interpreted by FB who, blinded for PET, identified the most plausible etiology if possible. This was meant to identify plausible evident etiologies that could not be identified by the majority algorithm. If still not possible, it was considered inconclusive.

> *[Comment from Prof. Dr. Frisoni: “I am not sure I agree that a consensus diagnosis based on clinical data and a 2-year follow-up is an appropriate reference. In patients with MCI, follow-up can elucidate converters from non-converters (although 2 years is a short span). In patients with dementia, the phenotype is not going to change in 2 years (and not even in 5 for that matter). AD phenotypes will stay AD and FTD phenotypes will stay FTD. Those AD who develop BPSD do not necessarily have FTD, and FTD who develop memory problems do not necessarily have AD. The notion of using a clinical phenotype as the reference dx goes against the modern notion of etiological diagnosis based on the molecular profile. I suggest to use the consensus dx only as a validation of the pre-PET dx by the local neurologist, not of the post-PET dx. This approach would IMHO save the dataset and the enormous amount of effort that went into the dx consensus (instead this is a strength of the dataset – if well exploited)*.*”]*

### STATISTICAL ANALYSIS

Demographic characteristics and the changes in etiology diagnosis from pre-PET to post-PET stratified by the reference etiology diagnosis were described.

For descriptive purposes the etiological diagnosis was classified as AD (including mixed AD) or non-AD. PET was considered to have had an added diagnostic value if the post-PET etiology diagnosis differed from the pre-PET etiology diagnosis and the post-PET etiology was in concordance with the reference etiology diagnosis. PET was considered to have generated a loss in diagnostic value if the post-PET etiology diagnosis differed from the pre-PET etiology diagnosis, and the post-PET etiology diagnosis was different from the reference etiology diagnosis. In all other situations, PET was considered to have no added diagnostic value and did not result in a loss in diagnostic value. Inconclusive reference outcomes were omitted from the analyses and described separately.

The proportion correct etiology diagnoses were tested both for pre-PET and post-PET using a McNemar’s chi-squared test using STATA15 (significance level set at 0.05). The power of the analysis was calculated using a power calculation for paired proportions, and minimal detectable difference under 80% power.

### SECONDARY AND SENSITIVITY ANALYSES

Several secondary and sensitivity analyses were performed to explore the data and asses the robustness of the results.

Sensitivity, specificity, positive predictive value (PPV), negative predictive value (NPV) was estimated for both the pre- and post-PET dichotomized etiology diagnosis in terms of AD versus non-AD.

The level of importance panelists rated the different categories of diagnostic information was described.

The association between baseline characteristics and correctness of the diagnostic change was estimated (i.e. to explore predicting in which individuals PET likely has an added value) using bivariate logistic regression analyses.

In a sensitivity analysis the McNemar’s test was performed using the certainty of the reference diagnosis as quasi weights.

The ratings from 3 panelists on etiology for those participants in which no majority etiology was reached and no plausible evident etiology could be set were listed.

The primary and part of the secondary analyses were performed both in- and excluding the plausible evident reference etiology diagnoses.

Due to the explorative nature of the sensitivity analyses, the significance levels were not adjusted for multiple testing.

## RESULTS

Part of the 1-year follow-up (n=4) and the 2-year follow-up (n=46) assessments were not performed leaving 102 participants with 2 follow-up and 50 participants with 1 follow-up assessment for analysis (see figure 1). Age, sex and education were similar, but MMSE (mean 23.8 versus 22.4), DAD (mean 85 versus 76) and NPI (mean 12 versus 19) were significantly more affected in the omitted part of the sample.

The average age was 62 years (range: 45-70) and 44% was female. The mean MMSE was 23.8 (SD=3.5). See table 1 for details.

**Table 1:** Sample characteristics at baseline by post-PET diagnosis (mean, SD or n, %).

|  | **AD (n=106)** | **FTD (n=18)** | **other NDD (n=18)** | **no NDD (n=10)** |
| --- | --- | --- | --- | --- |
| Age | 62 (6.1) | 62 (4.7) | 64 (5.3) | 60 (5.8) |
| Female gender | 51 (48%) | 7 (39%) | 6 (33%) | 3 (30%) |
| MMSE | 23 (3.2) | 24 (3.3) | 25 (4.3) | 25 (4.2) |
| DAD | 85 (14.8) | 80 (21.2) | 87 (12.3) | 88 (14.8) |
| GDS | 3 (2.3) | 3 (2.4) | 4 (2.9) | 3 (2.2) |
| post-PET diagnosis |  |  |  |  |
| AD | 86 (81%) | 3 (17%) | 2 (11%) | 5 (50%) |
| VAD | 1 (1%) |  | 3 (17%) |  |
| FTD | 4 (4%) | 15 (83%) |  |  |
| DLB | 2 (2%) |  | 6 (33%) |  |
| other NDD | 3 (3%) |  | 7 (39%) |  |
| no NDD | 10 (9%) |  |  | 5 (50%) |

Abbreviations: AD, Alzheimer's disease; DAD, disability assessment dementia (scale range 0-100, higher is less affected); DLB, dementia with Lewy bodies; FTD, frontotemporal dementia; GDS, geriatric depression scale (scale range 0-15, higher is more affected); MMSE, Mini-Mental State Examination (scale range 0-30, higher is less affected); NDD, neurodegenerative disease; VAD, vascular dementia.

A reference diagnosis was set in 122 participants (80%) with 56 a full and 66 a partly majority diagnosis (of whom 22 and 18 only had one of out of the 2 follow-up assessments available respectively). In 30 (20%) participants no majority was reached. Of these 30, in 22 a plausible evident etiology was set, leaving 8 participants with a definite non-majority etiology.

Of the subsample with full or partly majority diagnosis, in 107 (88%) cases post-PET was in line with the reference, and in 103 (84%) the pre-PET diagnosis was in line with the reference. The difference in proportions was 0.033 (95% CI -0.035 to 0.101; p=0.424). The power to detect this difference with the available sample size was 0.22 (a difference of 0.075 could be detected with the available sample size). Using the plausible evident etiology 122 post-PET (85%) and 118 pre-PET (82%) were in line with the reference (difference 0.028; 95% CI -0.037 to 0.092; p= 0.481; power=0.16; detectable difference with sample size was 0.077).

In 20 cases (16%) the diagnosis changed between pre-PET to post-PET. Of those 20, 9 changes were correct (i.e. post-PET in line with reference), 7 changes were incorrect (i.e. post-PET differed from reference), 4 changes were within a mixed reference etiology (e.g. pre-PET AD to post-PET FTD, majority etiology was mixed AD/FTD). When including the plausible evident cases, 27 (19%) changes occurred of which 11 correct, 11 incorrect and 5 within mixed.

See table 2 for the details stratified by the reference diagnosis.

**Table 2:** pre-PET diagnosis versus post-PET diagnosis, stratified by the reference diagnosis.

| Pre-PET | Post-PET | Reference diagnosis | Pre/post-PET change? | If change, in line with reference? | Regardless of change, post-PET in line with reference? | Comment | n<sup>1</sup> | n<sup>2</sup> |  |
| --- | --- | --- | --- | --- | --- | --- | --- | --- | --- |
| *CORRECT CHANGE* |  |  |  |  |  |  |  |  |  |
| noAD | AD | AD | Yes | Yes | Yes |  | 5 | 7 | A |
| AD | noAD | noAD | Yes | Yes | Yes |  | 3 | 3 | B |
| AD | noAD | mixed noAD | Yes |  | Yes | post-PET within mixed | 1 | 1 | C |
| *INCORRECT CHANGE* |  |  |  |  |  |  |  |  |  |
| AD | noAD | AD | Yes | No | No |  | 4 | 5 | D |
| AD | noAD | mixed AD | Yes | No | No | post-PET outside mixed | 1 | 2 | E |
| AD | noAD | noAD | Yes | No | No | post-PET different noAD than reference noAD | 2 | 4 | F |
| *CHANGE WITHIN MIXED* |  |  |  |  |  |  |  |  |  |
| AD | noAD | mixed AD | Yes |  | Yes | post-PET within mixed | 4 | 5 | H |
| *NO CHANGE (INCORRECT)* |  |  |  |  |  |  |  |  |  |
| AD | AD | noAD | No | n/a | No |  | 2 | 3 | N |
| noAD | Same noAD | AD | No | n/a | No |  | 2 | 3 | O |
| noAD | Same noAD | noAD | No | n/a | No | post-PET different noAD than reference noAD | 4 | 5 | P |
| *NO CHANGE (CORRECT)* |  |  |  |  |  |  |  |  |  |
| AD | AD | AD | No | n/a | Yes |  | 65 | 70 | I |
| AD | AD | mixed AD | No | n/a | Yes |  | 11 | 12 | J |
| noAD | same noAD | noAD | No | n/a | Yes |  | 13 | 18 | K |
| noAD | same noAD | mixed AD | No | n/a | Yes | post-PET within mixed | 3 | 4 | L |
| noAD | same noAD | mixed noAD | No | n/a | Yes | post-PET within mixed | 2 | 2 | M |
| *NO CONSENSUS* |  |  |  |  |  |  |  |  |  |
| AD | AD | no consensus | n/a | n/a | n/a |  | 8 | 1 | Q |
| AD | noAD | no consensus | n/a | n/a | n/a |  | 5 | 1 | R |
| noAD | AD | no consensus | n/a | n/a | n/a |  | 5 | 2 | S |
| noAD | same noAD | no consensus | n/a | n/a | n/a |  | 11 | 3 | T |
| noAD | other noAD | no consensus | n/a | n/a | n/a |  | 1 | 1 | U |
| *Total* |  |  |  |  |  |  | 152 | 152 |  |

<sup>1</sup> Excluding plausible evident cases.
<sup>2</sup> Including plausible evident cases.

[Alternative table for consideration]
| Pre-PET | Post-PET | Change in clinical diagnosis after PET? | correct change according to reference standard | incorrect change according to reference standard | change within mixed diagnosis set in reference standard | no change (diagnosis different from reference diagnosis) | no change (diagnosis same as reference diagnosis) | no consensus |
| --- | --- | --- | --- | --- | --- | --- | --- | --- |
| noAD | AD | Yes | 5 |  |  |  |  | 5 |
| AD | noAD | Yes | 4 | 7 | 4 |  |  | 5 |
| AD | AD | No |  |  |  | 2 | 76 | 8 |
| noAD | same noAD | No |  |  |  | 6 | 18 | 11 |
| noAD | other noAD | Yes |  |  |  |  |  | 1 |

Figure 2 displays the pre- and post-PET diagnostic certainty classified by correct, incorrect or no change in etiological diagnosis. In a large proportion of the participants the diagnostic certainty increased.

**Figure 2:**
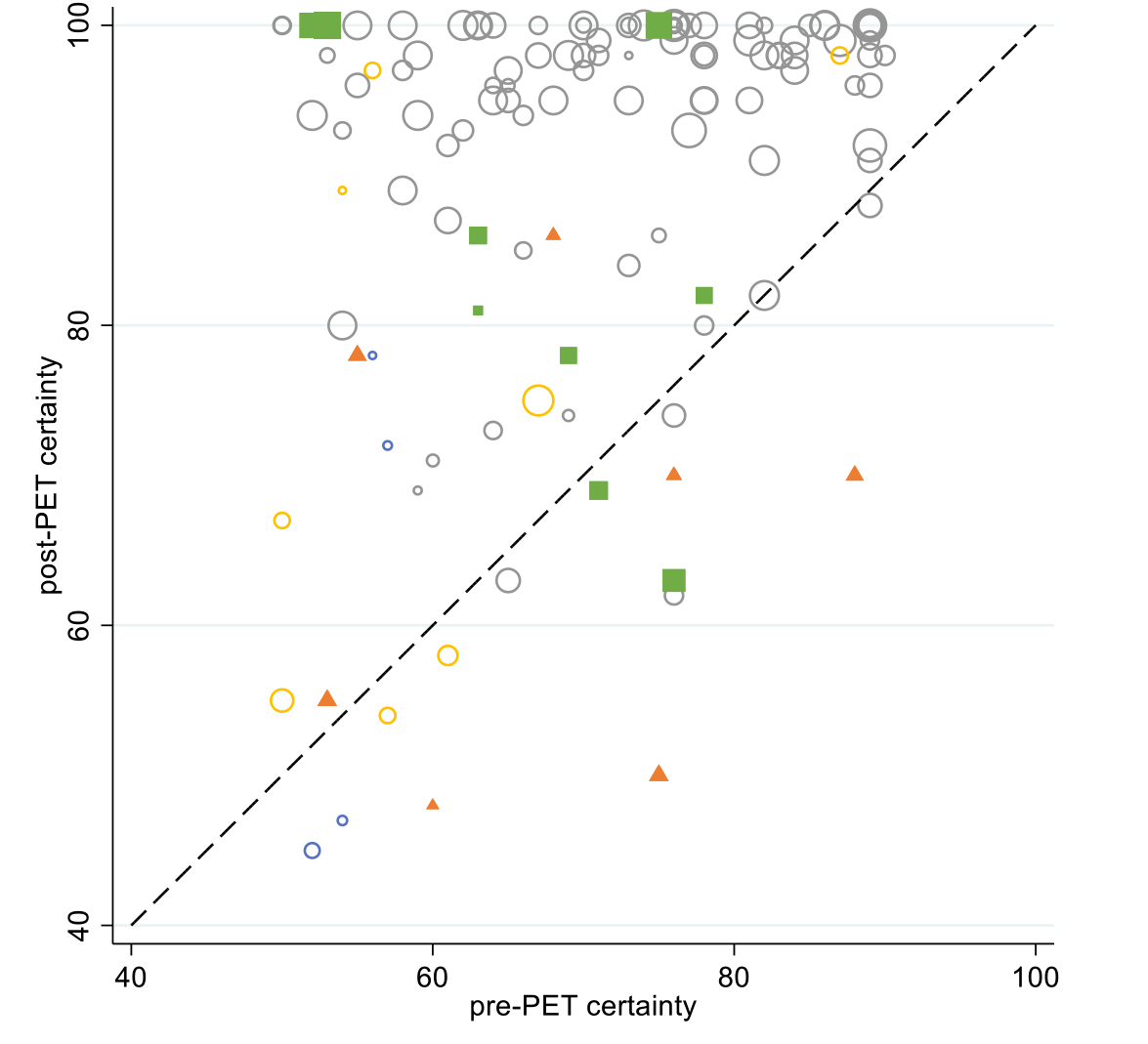
Pre- and post-PET certainty of each participant classified according to table 2: green filled squares: change concordant with reference standard, orange filled triangles: change discordant with reference standard, blue open circles: changed within mixed, yellow open circles: no change and concordant to reference standard, gray open circles: no change and concordant to reference standard; dashed black line is diagonal; circle size is reflection of the relative certainty within each of the 5 classifications.

### SECONDARY AND SENSITIVITY ANALYSES

Pre-PET sensitivity and specificity were 0.89 and 0.70 and post-PET were 0.85 and 0.92 respectively. Sensitivity did not significantly change, while specificity did significantly change (p=0.031). This was 0.86 and 0.69 pre-PET, and 0.83 and 0.92 post-PET respectively when including plausible evident etiology ratings, also with significant specificity change.

PPV and NPV was 0.91 and 0.66 for pre-PET and 0.98 and 0.64 for post-PET respectively. This was 0.89 and 0.63 for pre-PET, and 0.97 and 0.65 for post-PET when including plausible evident etiology ratings.

The mean (standard deviation) level of importance ratings from the panelists of the different categories of diagnostic information on a scale from 1 to 15 was 11.1 (2.5) for clinical information, 9.9 (4.7) for CSF, 10.1 (3.0) for MRI, 9.8 (2.7) for neuropsychiatric examination and 5.4 (5.7) for other information (such as blood test or FDG PET).

Bivariate logistic regression on correct versus incorrect or no change (with change within mixed classified as no change) using age, gender, education, MMSE, CDR, GDS, DAD, dichotomized pre-PET diagnosis (AD/non-AD) and pre-PET diagnostic certainty indicated only pre-PET diagnosis not being AD was significantly associated to a correct change after PET (both without and with the plausible evident cases). An alternative analysis classifying change within mixed as change indicated lower CDR and lower pre-PET certainty were significantly associated to this (and in addition to these 2 also pre-PET diagnosis not being AD when using plausible evident cases).

The quasi-weighted McNemar’s test indicated the proportion correctly diagnoses was not significantly different.

Supplemental 5 describes the ratings from 3 panelists on etiology for those participants in which the no majority etiology diagnosis was reached and no plausible evident etiology diagnosis could be set.

## DISCUSSION

In a cohort of 122 young onset dementia patients in an academic memory clinic [18F]flutemetamol PET performed after the usual diagnostic workup resulted in a correct change of the etiological diagnosis in 9 participants and an incorrect change in 7 participants, as compared to a panel reference etiology using up to 2 year follow-up information on clinical symptoms. The percentage correctly diagnosed was 84% pre-PET and 88% post-PET (not significant).

In this study [18F]flutemetamol PET did not result in a significant improvement of correct diagnostic changes. However, the statistical power was insufficient to establish the significance of the relatively small difference that was observed (4% improvement). An explanation why PET did not result in a larger improvement is that the standard diagnostic workup already includes an extensive set of diagnostic information on patient history, and physical, neurological, psychiatric, neuropsychological and MRI. CSF was obtained in part of the sample (xx%), but the rated the pre- and post-PET diagnosis were rated blinded for CSF results. The added value of PET was possibly further limited if CSF would have been part of the pre-PET diagnostic workup, as amyloid PET likely is associated to CSF amyloid beta.

In addition, mean pre-PET certainty on the etiology was 70 (on a scale of 1-100; SD=12, and the highest quartile ranged from 79 to 89), which could be considered relatively high. Post-hoc analyses on a subsample of those with 50 to 65 pre-PET uncertainty indicated the percentage correctly diagnosed was 71% in pre-PET and 79% in post-PET. This indicates a possible higher value in more uncertain cases. The secondary analysis (including the plausible evident cases) further support this as it indicated that pre-PET diagnosis not being AD, a lower pre-PET certainty as well as less affected cognition and function (measured by the CDR) were associated to improved correct diagnostic changes. The less affected symptoms might indicate a possible added value in persons with very mild dementia although this was not confirmed by the secondary analysis on function measured by the DAD and cognition measured by the MMSE. The possible higher added value in more uncertain cases is further supported by the findings of Hellwig et al. [2018] who assessed a sample of patients with a complicated clinical presentation. They found a lower pre-PET accuracy of 71% (compared to 84% in our study) and a larger proportion of 23% in whom the diagnosis was changed (compared to 16% in our study), arriving at a post-PET accuracy of 89% (compared to 88% in our study). The plausibility that PET has a larger added value in uncertain cases is further supported by the study by Ceccaldi et al. [2018] who reported a large change in diagnosis (67%) in a sample of patients in whom CSF was not possible or even after CSF testing the etiology remained uncertain. However, opposite to the study by Hellwig et al., this study did not assess the accuracy by means of a (panel) reference standard. Figure 2 is also in line with this explanation as it shows only 1 diagnostic change in those with a pre-PET certainty of 80 or higher, reflecting no added diagnostic accuracy value (other than the potential value of increased certainty).

The vignettes used in the reference standard were composed of both clinical symptoms (phenotypical expression) as well as pathological measures (i.e. MRI and CSF), although longitudinal information on pathological measures was only available in a few cases.

Therefore, the [18F]flutemetamol PET scan result could have correctly identified amyloid pathology while being discordant with a non-AD reference outcome. Postmortem studies have shown AD and DLB or AD pathology can be both present [Harding, 2001], and clinical expressions of FTD with AD pathology present [Johnson, 1999]. Although this seems contradicting, it is in line with the purpose of this study to assess the clinical utility of PET (rather than its correlation to pathology) as clinical management is oriented towards current but also future symptoms, such as practical support in home setting, treatment and case management.

Obviously in case of future anti-amyloid therapy it becomes essential to assess PET in terms of its association to pathology, which has indicated a relatively high accuracy [Curtis, 2015].

Alternatively, the [18F]flutemetamol PET result could have been included in the vignettes for the reference standard, as has been done by Hellwig et al. [2018]. This could have improved the reference standard, especially for atypical representations. It would likely have resulted in a different reference diagnosis in the 8 cases with ‘no change (incorrect)’ (see table 2).

However, its importance could be overestimated causing incorporation bias and leading to an overestimation if the PET’s accuracy [whiting, 2004] and therefore often advised not to [Rutjes, 2007].

### STRENGTHS AND LIMITATIONS

This study relied on a reference standard based on longitudinal data, which generated empirical evidence in a relatively large sample of 122 participants on the correctness of the diagnostic changes due to [18F]flutemetamol PET in an academic setting.

Our study was limited due to several reasons. Drop-out was relatively large (28%) and diagnostic changes occurred less in the sample remaining for analysis (16% instead of 19% in the total sample [Zwan, 2017]). This could have resulted in a drop-out bias underestimating the clinical utility of [18F]flutemetamol PET. However, of the 3 factors associated with a correct diagnostic change (CDR, pre-PET diagnostic certainty and pre-PET diagnosis of AD) only CDR was higher in the omitted sample (0.84 versus 0.71 in the sample remaining for analysis), which makes a large underestimation unlikely.

We have altered usual practice by excluding CSF from the pre- and post-PET diagnosis. It is expected that if amyloid beta CSF was included in the pre-PET diagnosis, it would have reduced the added diagnostic value of PET due these both amyloid tests being correlated. Nevertheless, the current estimates reflect the effect of PET if a lumbar puncture could not be performed due to for example contra-indications, technical problems or patient refusal.

The follow-up period of 2 years possibly limited a confident reference diagnosis in participants with slow disease progression as for example AD pathology could come to expression after 2 years (i.e. imperfect reference standard bias). However, a too long follow-up up to a decade would drive the reference standard towards a pure prognostic marker as it becomes less likely that longer-term symptoms retrospectively apply to the baseline situation (i.e. disease progression bias: when index test is performed unusually long before the reference standard, such that the disease is more advanced when the reference standard is performed).

A panel reference standard requires a large time investment of panelists, and would be even larger if a stepwise evaluation was incorporated (e.g. to assess the value of including PET in the reference standard by subsequently providing [18F]flutemetamol PET results in the patient vignettes). Alternative to a panel reference standard a composite or latent reference standard could be determined based on all information from the vignette [Bertens, 2013]. However, this would omit the information from clinical history and is complex as the vignette includes a large variety of detailed information. Due to time constraints some recommendations were omitted [Bertens, 2013] possibly limiting the validity of the study: including 3 panelists instead of 5 or 7, no validation assessment in a subsample, no video recordings to see the participant history interview ‘live’, and no (staged) discussion meeting to reach consensus on non-majority cases [Bertens, 2014]. As an alternative to the latter, the panelists ratings of each of the 8 non-majority cases were assessed by a single clinician (FB, blinded for PET and not being one of the panelists). As the analyses including these data showed similar results to the analyses excluding this approach showed similar results, the conclusion is likely unaffected by this design choice. In addition, we believe that a forced consensus on cases with large disagreement among panelists likely results in arbitrary reference diagnoses. However, these are probably important cases as the proportion of change was larger in the 8 cases without a reference etiology diagnosis available (50%) than in the 122 cases with a reference etiology diagnosis available (16%).

### CLINICAL AND RESEARCH IMPLICATIONS

Based on the primary outcome of no increased significant diagnostic accuracy our study does not support the implementation of [18F]flutemetamol amyloid PET in routine clinical academic practice.

However, based on the secondary and post-hoc analyses we hypothesize a benefit of amyloid PET (in terms of increased diagnostic accuracy when compared to a reference standard) in academic clinical practice in persons with low pre-PET diagnostic certainty.

Diagnostic accuracy remains an intermediate outcome for the final goal of improving patients’ health or limit health loss. Therefore, future studies should focus beyond this concept and assess health outcomes even in absence of disease-modifying treatments [Bossuyt, 2009], preferably in a randomized setting with and without (disclosing) amyloid PET [Lijmer, 2009b].

### CONCLUSION

[18F]flutemetamol amyloid PET did not significantly improve the diagnostic accuracy in young patients with dementia in an academic memory clinic. The secondary analyses indicated the potential added value of amyloid PET in a narrower subsample of persons with (very) high diagnostic uncertainty, which we recommend to assess in future research.

## Data Availability

All data produced in the present study are available upon reasonable request to the authors

## ACKNOWLEDGMENT

This study was performed within the framework of the Dutch Flutemetamol Study and supported by the Dutch Alzheimer’s Society (grant WE.15-2014-01) and through an unrestricted grant of GE Healthcare to the Stichting Alzheimer & Neuropsychiatrie, Amsterdam. Research of the VUmc Alzheimer Center is part of the neurodegeneration research program of the Neuroscience Campus Amsterdam. The VUmc Alzheimer Center is supported by Alzheimer Nederland and Stichting VUmc fonds. The clinical database structure was developed with funding from Stichting Dioraphte.

Dutch Flutemetamol Study EudraCT Number 2012-002303-18 (clinicaltrialsregister.eu).

Prof. Dr. Frisoni has provided written consent to publish his comment and name as provided in this manuscript.

## SUPPLEMENTAL 1: IN- AND EXCLUSION CRITERIA

### INCLUSION CRITERIA

Patients:

- Who are aged less than 70 at the moment of the diagnosis;
- Who have dementia or are suspected of dementia;
- Whose etiological diagnosis was set with a certainty between 50-90%;
- Who underwent a brain MRI during a standardized screening;
- Who signed an informed consent prior to the study performances;
- Who are mentally competent (this was operationalized by a Mini Mental State Examination score ≥ 18);
- Whose body weight is more or equal to 50 kilos (this was done due to PET scanning properties).

### EXCLUSION CRITERIA

Patients:

- Who are considered medically unstable;
- Who require additional laboratory tests or workup between enrolment and completion of the [18F]flutemetamol PET scan;
- Who have a clinically significant infectious disease, including Acquired Immunodeficiency Syndrome (AIDS) or Human Immunodeficiency Virus (HIV) infection;
- Who are receiving any investigational medications, or have participated in a trial with investigational medications within the last 30 days;
- Who have ever participated in an experimental study with an amyloid targeting agent (e.g. anti-amyloid immunotherapy, γ-secretase or γ-secretase inhibitor) unless it can be documented that the subject received only placebo during the course of the trial;
- Who have had a radiopharmaceutical imaging or treatment procedure within 7 days prior to the study imaging session;
- Who are females of childbearing potential who are not surgically sterile, not refraining from sexual activity or not using reliable methods of contraception. Females of childbearing potential must not be pregnant (negative serum β-hCG at the time of screening and negative urine β-hCG on the day of imaging) or breast feeding at screening. Females must avoid becoming pregnant, and must agree to refrain from sexual activity or to use reliable contraceptive methods such as prescribed birth control or IUD for 24 hours following administration of [18F]flutemetamol;
- Who are claustrophobic;
- Who have abnormalities on MRI other than white matter changes or an incidental small lacunar lesion;
- Who have metal objects in or around the body (braces, pacemaker, metal fragments).

## SUPPLEMENTAL 2: EXAMPLE VIGNET WITH HYPOTHETICAL DIAGNOSTIC INFORMATION

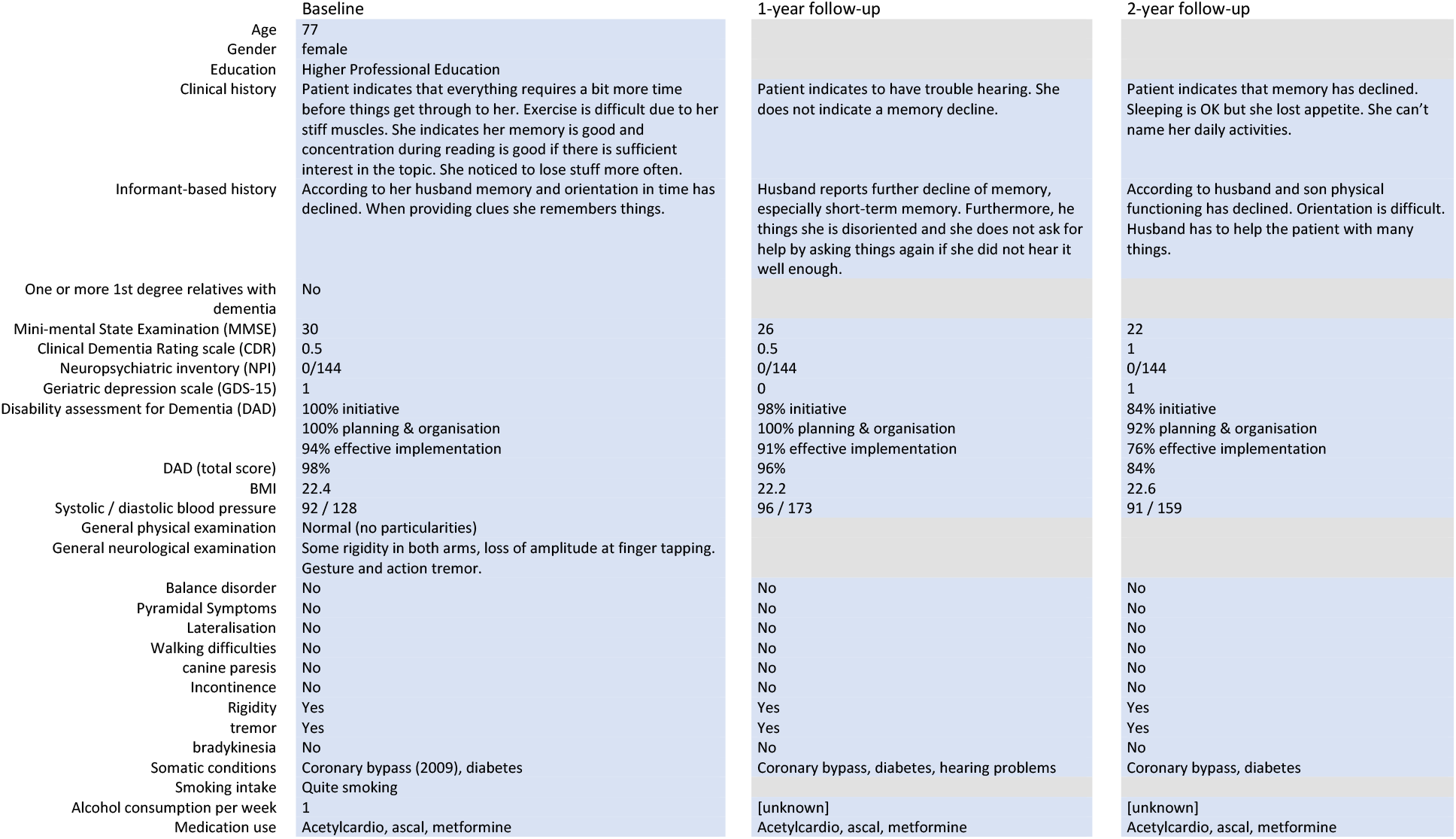
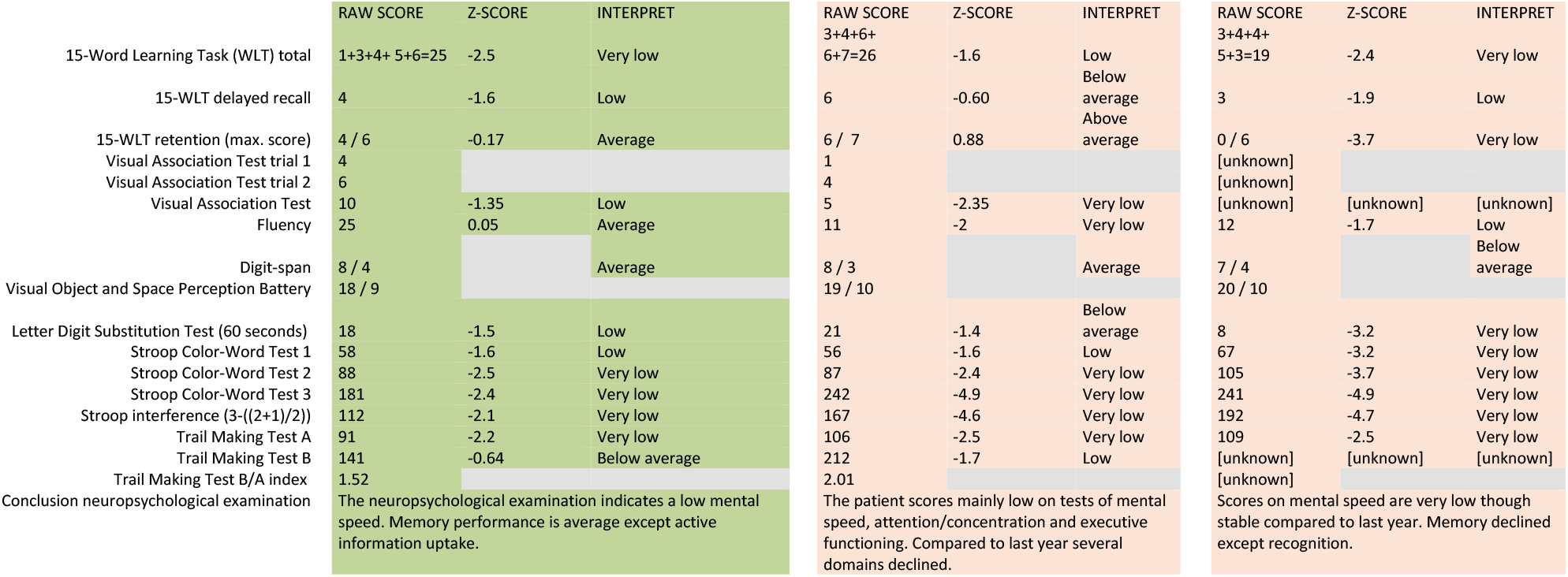
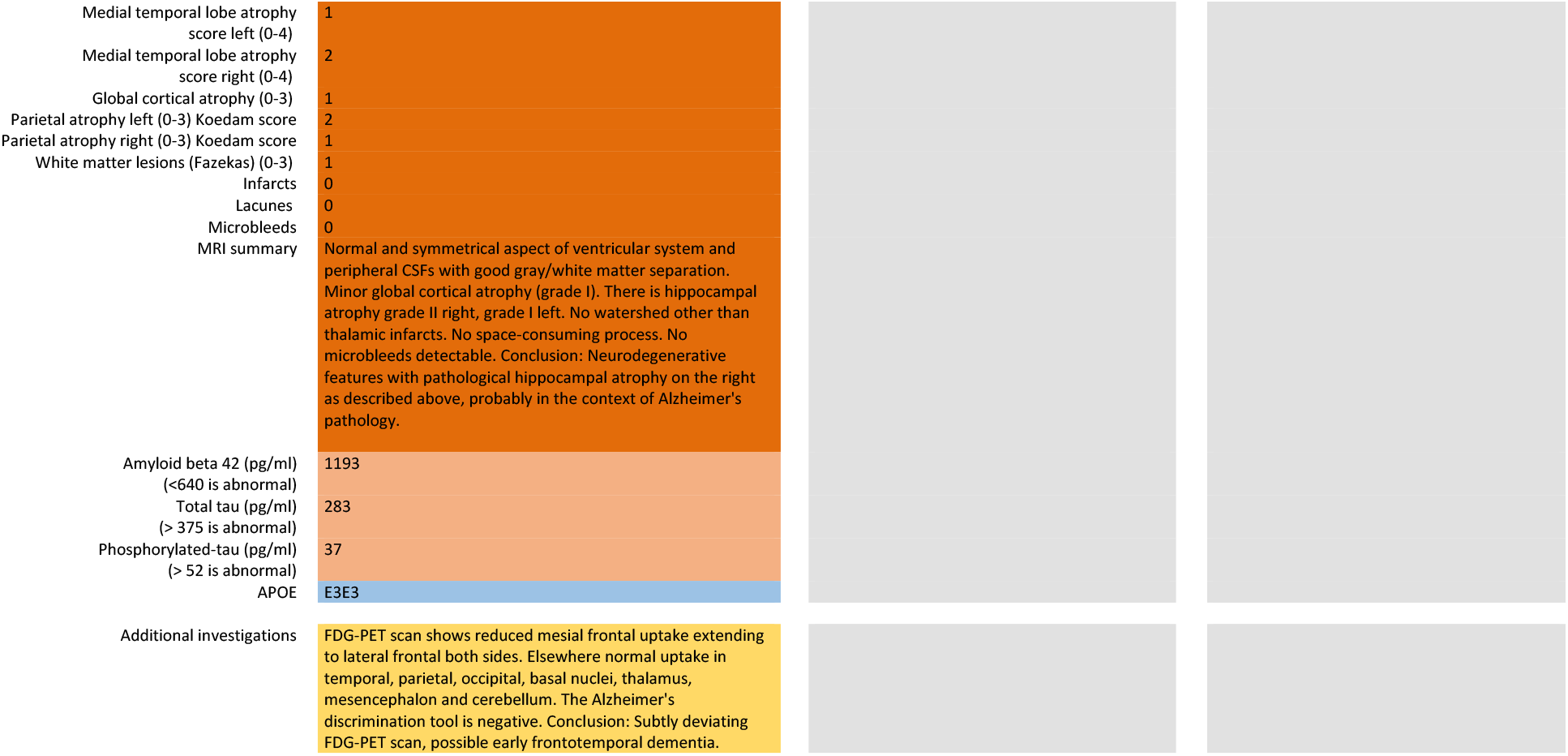

## SUPPLEMENTAL 3: DIAGNOSTIC ASSESSMENTS

### PRE-PET DIAGNOSIS

Item 1: Most likely etiology (choose 1 option):

- Alzheimer
- Vascular
- Frontotemporal
- Lewy Bodies
- Other neurodegenerative condition, namely
- No neurodegenerative condition, namely
- Unclear / postponed

Item 2: Degree of certainty on most likely etiology (mark on the line):

Completely uncertain |__________________________| Completely certain

Item 3: Etiological differential diagnosis (choose 1 option):

- Alzheimer
- Vascular
- Frontotemporal
- Lewy Bodies
- Other neurodegenerative condition, namely________________
- No neurodegenerative condition, namely______________

### POST-PET DIAGNOSIS

Item 1: Most likely etiology (choose 1 option):

- Alzheimer
- Vascular
- Frontotemporal
- Lewy Bodies
- Other neurodegenerative condition, namely___________
- No neurodegenerative condition, namely__________
- Unclear / postponed

Item 2: Degree of certainty on most likely etiology (mark on the line):

Completely uncertain |--------------------------------------------------------| Completely certain

### REFERENCE DIAGNOSIS

<u>Item 1: Which etiology determines the clinical picture?</u>

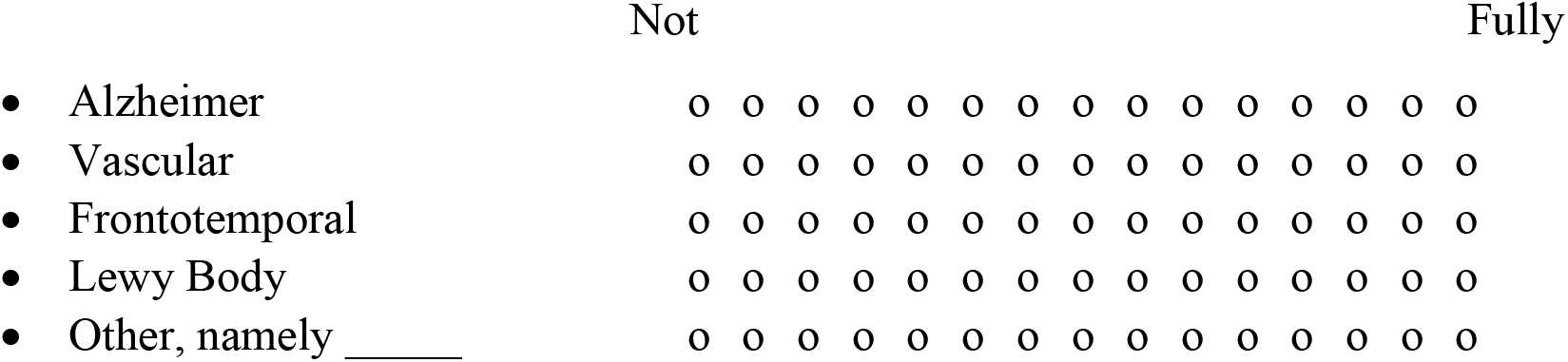

<u>Item 2: How certain are you of this etiology?</u>

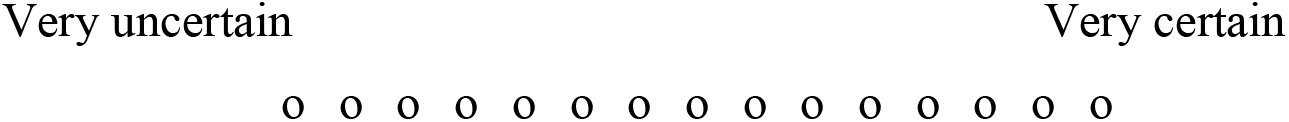

<u>Item 3: How much value do you attach to the different parts of information to determine the etiology?</u>

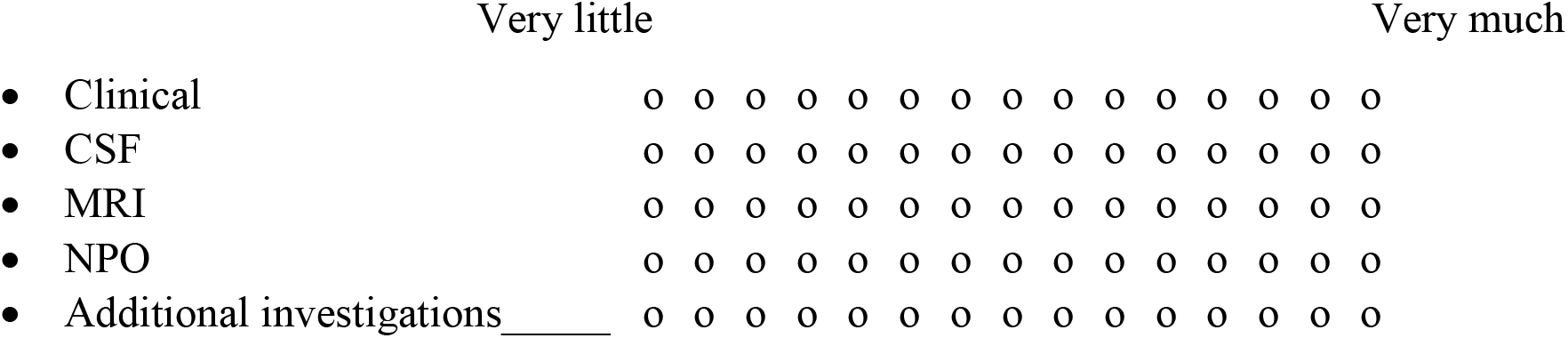

## SUPPLEMENTAL 4: EXAMPLES OF MAJORITY

### EXAMPLE 1

*Individual expert’s scores (on scale of 1 to 15):*

Expert 1: 7 for AD and 3 for VAD.

Expert 2: 11 for AD and 7 for VAD.

Expert 3: 9 for AD, 4 for VAD and 2 for other NDD.

*The weighted scores are:*

Expert 1: AD: 7/(7+3) = 0.70; VAD: 3/(7+3) = 0.30 Expert 2: AD: 11/(11+7) = 0.61; VAD: 7/(11+7) = 0.39

Expert 3: AD = 10/(10+3+2) = 0.67; VAD: 4/(10+3+2) = 0.20; other NDD 2/(10+3+2) = 0.13

*Diagnoses with weight higher than or equal to 0*.*33:*

Expert 1: AD

Expert 2: AD, VAD

Expert 3: AD

*Majority diagnosis:*

AD

### EXAMPLE 2

*Individual expert’s scores:*

Expert 1: 7 for AD and 3 for VAD (on scale of 1 to 15).

Expert 2: 11 for AD and 7 for VAD.

Expert 3: 9 for AD and 4 for VAD.

*The weighted scores are:*

Expert 1: AD: 7/(7+3) = 0.70, VAD: 3/(7+3) = 0.30

Expert 2: AD: 11/(11+7) = 0.61, VAD: 7/(11+7) = 0.39

Expert 3: AD = 9/(9+4) = 0.69, VAD: 4/(9+4) = 0.31

*Diagnoses with weight higher than or equal to 0*.*33:*

Expert 1: AD

Expert 2: AD, VAD

Expert 3: AD

*Majority diagnosis:*

AD

## SUPPLEMENTAL 5: NO (PLAUSIBLE) MAJORITY

Ratings from 3 experts on etiology (“Which etiology determines the clinical picture”; scale from 1 to 15) for those 8 participants in which the no majority diagnosis was reached on the etiology and no plausible evident etiology could be set.

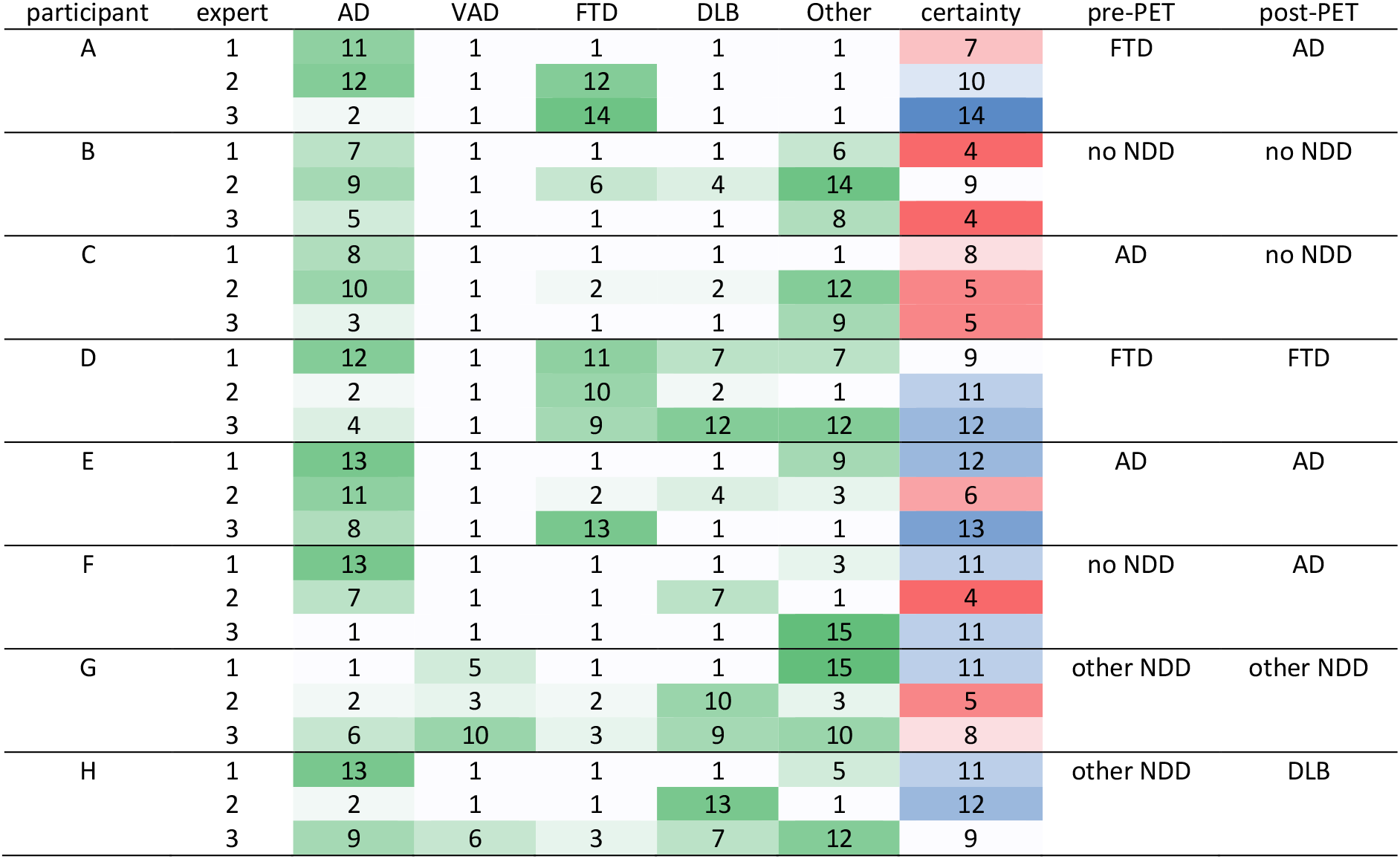

